# Serum Vitamin D Levels do not Affect Susceptibility to EBV Infection – A Mendelian Randomization Study

**DOI:** 10.64898/2026.10.29.26366817

**Authors:** Jack Stanley, George Davey Smith, Fergus Hamilton

## Abstract

**Introduction:** Epstein–Barr virus (EBV) infection and low serum 25-hydroxyvitamin D (25OHD) levels are strongly associated with multiple sclerosis (MS). We investigated whether genetically proxied serum vitamin D levels are causally linked to EBV infection, assessed by serology and detectable EBV DNA, using Mendelian randomisation (MR).

**Methods:** We conducted two-sample cis-MR using genetic instruments associated with serum 25OHD concentrations, restricted to variants near genes implicated in vitamin D metabolism. Outcomes included EBV seropositivity, antibody responses to four EBV antigens, detectable EBV DNA in blood and saliva, and MS case status as a positive control.

**Results:** Genetically proxied vitamin D levels had no clear effect on EBV seropositivity (odds ratio per standard deviation (SD) increase in rank-based inverse-normal transformation (RINT) 25OHD: 1.76, 95% confidence interval [CI] 0.83–3.73) or antibody responses to EBV antigens. No significant associations were observed with detectable EBV DNA in blood (beta 0.02, 95% CI −0.004 to 0.04) or saliva (beta 0.02, 95% CI −0.23 to 0.26). In contrast, genetically proxied vitamin D levels were associated with reduced MS risk (odds ratio per SD increase in RINT 25OHD 0.67, 95% CI 0.54–0.84).

**Discussion:** We found no clear evidence that genetically proxied vitamin D levels causally influence EBV serological responses or detectable EBV DNA in blood or saliva. The protective association with MS supports previous genetic evidence. These findings suggest that vitamin D and EBV may influence MS risk through distinct pathways, although effects specific to early-life vitamin D exposure remain possible.

## Introduction

Vitamin D is a fat-soluble vitamin and steroid pro-hormone that serves as the precursor to 1,25-dihydroxyvitamin D (1,25(OH)_2_D or calcitriol)(1), which is measurable in serum providing a measure of vitamin D status. A number of genetic variants have been shown to influence vitamin D levels(2) with the largest recent GWAS highlighting 143 SNPS which explain ∼13% of variation in vitamin D levels.

Vitamin D has direct effects upon calcium and phosphate metabolism, impacting bone health. Severe vitamin D deficiency causes nutritional rickets and decreased bone mineral density(3). Vitamin D receptors (VDR) are however widely distributed throughout the human body(4) and vitamin D itself acts as a precursor to transcription factors acting on genes implicated in protection against carcinogenesis and immune function.

Observational studies suggested extra-skeletal effects for vitamin D on conditions including cancer, ischaemic heart disease and response to infection(5, 6). However subsequent large randomized-controlled trials and meta-analyses failed to substantiate these findings, suggesting observations were related to reverse causality or residual confounding(7-9).

Mendelian randomization (MR) is a methodological approach to causal inference which can minimize bias related to residual confounding, reverse causality, and exposure misclassification. MR has been applied to a number of observed and hypothesized vitamin D-health outcome associations(10). Yet the use of the approach in this field is not without its own challenges(11). These include use of non-linear MR (NLMR) approaches which have led to spurious reporting of causal relationships between vitamin D levels and cardiovascular and mortality outcomes, necessitating retraction(11, 12) MR analysis using reliable methods and appropriately constrained (ie using only variants on the vitamin D pathway), cis-MR analyses of vitamin D have failed to show any extra-skeletal effect beyond a strong inverse effect on the risk of multiple sclerosis (MS)(10, 13).

MS is a complex trait which develops in genetically susceptible individuals exposed to environmental stressors. Two closely associated environmental stressors, first highlighted in migration studies and repeatedly reinforced are low vitamin D levels, which are one postulated explanation for the increasing incidence of MS with increasing latitude(14) and infection with EBV (Epstein-Barr virus) at a specific stage of the life course(15, 16).

Elevated levels of antibodies to EBNA (Epstein-Barr Nuclear Antigen) have been found to predate the onset of MS by up to 20 years(17) and the brains of those with MS, and particularly the lesions characterising MS itself, have been shown to more commonly demonstrate evidence of EBV infection than the brains of individuals unaffected by MS(18). Furthermore variation in DRB1*15:01 (a Human Leukocyte Antigen locus on chromosome 6) has been identified as a key genetic risk factor for development of MS and has also been associated with EBV ZEBRA (BamHI Z Epstein-Barr virus replication activator) antibody levels(19).

Given the high prevalence of EBV infection in the general population (adult seroprevalence exceeds 90% worldwide(20)) in comparison to the relatively lower population prevalence of MS (35.9/100,000 worldwide(21)) it is clear that EBV infection alone is not enough to cause MS. Host specific factors influencing immune control, including but not limited to the age of EBV acquisition are hypothesized to explain why only certain individuals go on to develop MS and other post-infective sequelae(20, 22).

In most cases acute EBV infection is followed by lifelong viral persistence in dormant form in B lymphocytes, as well as the epithelial cells of the oropharynx. In these individuals serological testing demonstrates the host immune response to primary infection(23). In a subset of cases immune control of the infection is lost and EBV DNA can be detected in the peripheral blood. A recent genome-wide association study (GWAS) exploited the fact that whole genome sequencing (WGS) of human peripheral blood detects EBV DNA if it is present. This measurement reflects detectable EBV DNA at a single time-point, reflecting a snapshot of a dynamic process, which is known to fluctuate temporally within individuals(24). They extracted all sequencing reads from UK Biobank (UKB) and All of Us (AoU) that mapped to the EBV genome to quantify EBV DNA coverage and elucidate single nucleotide polymorphisms (SNPs) causally linked to detectable EBV DNA (under the sequencing conditions used by UKB and AoU) (25). This work was expanded upon by a subsequent paper which utilised a similar approach in both blood and saliva, showing that the presence of detectable EBV in saliva (∼38% prevalence with at least one EBV DNA read pair) is more common than in blood (∼15% prevalence with at least one EBV DNA read pair)(26). It is hypothesized that salivary detection represents evidence of active (lytic) viral replication but does not necessarily indicate active viraemia and as such detection of EBV DNA in saliva and blood represent different clinical phenotypes of virological control(26).

Vitamin D is known to interact with broad ranging cells across the body, including those involved in the immune system where it has myriad effects, including suppression of B lymphocyte differentiation(27). We hypothesize that vitamin D levels may influence immune control of EBV and explain the as-yet unexplained effect of low vitamin D on the risk of MS.

We aim to investigate, using mendelian randomization (MR)(28-30), whether serum vitamin D levels are causally linked to a) serological evidence of EBV infection and b) detectable EBV DNA in blood and saliva.

## Methods

### Study design and data sources

The analytic approach was two sample *cis* Mendelian randomisation(29). This approach exploits the natural genetic variation and utilises single nucleotide polymorphisms (SNPs) that associate with an exposure to estimate the causal effect of the exposure on an outcome.

We used a recent, multi-ancestry GWAS (genome wide association study) (n = 417,580 participants in UK Biobank) reporting vitamin D as rank-based inverse-normal transformation (RINT) 25OHD(2) to extract instrumental variables (IVs) for our analysis (Supplementary Table 1). In this GWAS individuals had total 25OHD (combined 25OHD_3_ and 25OHD_2_) measured using the Diasorin Liason®, a chemiluminescent immunoassay (CLIA).

### Assumptions about instruments

To satisfy the three core assumptions for MR in an IV framework(28) of relevance, independence and exclusion restriction we applied the following criteria to our IVs. We restricted instruments to those that are cis (within 10^6^ base pairs) to genes known to be on the vitamin D pathway (*CYP2R1, GC, NADSYN1/ DHCR7, CYP24A1*), independent (R^2^ > 0.1) and strongly associated with vitamin D blood levels (p < 5 x 10^-8^). Furthermore, we assessed for horizontal pleiotropy as described below.

### Outcomes

As outcomes we analysed a) EBV serological markers, b) presence of EBV DNA in participants blood (EBV DNAemia) and saliva and c) diagnosis of MS with the latter included as a positive control in light of the extensive evidence from genetic studies around the effect of vitamin D levels on MS(10, 13). The studies used are described in Supplementary Table 1 and expanded below.

For EBV serology we used data from a study of 9724 participants who provided serum samples for serological measurements to analyse seropositivity for EBV (positive for 2 or more antigens, n = 8477, 94.4% positive), median fluorescence intensity (MFI) levels of VCA p18 (Viral Capsid Antigen protein 18)(n=8518, 94.8% seropositive), EBNA-1 (n=7972, 88.7% positive), ZEBRA (n = 8191, 91.2% positive), EA-D (Early Antigen-Diffuse) (n = 7763, 86.4% positive)(19).

For detectable EBV DNA we used a recent GWAS analysing blood and salivary detection of EBV DNA, reporting inverse normal transformed (INT) counts of EBV DNA read pairs (Supplementary Table 1) (26). The R package MungeSumstats was used to format data from these GWAS(31).

For MS we used a 2019 GWAS which analysed 115,803 individuals including 47,429 cases(32) diagnosed by practicing neurologists according to the clinical guidance in the country of diagnosis.

### Statistical methods: main analysis

Estimates were generated by the Wald Ratio and analysed by inverse-variance weighting (IVW). Coefficients were exponentiated to generate odds ratios (OR) for binary outcomes. For non-binary outcomes the effect estimate (beta) was reported on the original scale. In both cases 95% confidence intervals (95% CI) were reported. The analysis was performed in R version 4.5.3(33) using the Two-Sample MR package(34). Exposure-associated SNPs that were missing in the outcome dataset were excluded from the analysis.

### Statistical methods: sensitivity analysis

We repeated our IVW MR with IVs as described above, including the requirements for independence and strong association with vitamin D levels but omitting the requirement for the SNPs to be cis to genes known to be on the Vitamin D pathway.

We also report the results of MR using Egger, Simple Mode, Weighted Mode and Weighted Median as sensitivity analyses. To assess for horizontal pleiotropy we performed MR Egger and report the p value and intercept. To assess for heterogeneity we report Cochran’s Q for the IVW analysis.

### Ethics

No formal ethics approval was needed for this study as publicly available data was used. This study was not pre-registered.

## Results

We had 17 SNPs as IVs, summarised in Supplementary Table 1. SNP level data is provided in Supplementary Tables 2 and 3.

### EBV serology

There is clear evidence of a protective effect of higher levels of vitamin D on multiple sclerosis case status (Figure 1) (Odds ratio (OR) per standard deviation (SD) change in (RINT) vitamin D 25OH-D levels 0.67 (95% confidence interval (CI) 0.54-0.84).

**Figure 1.**
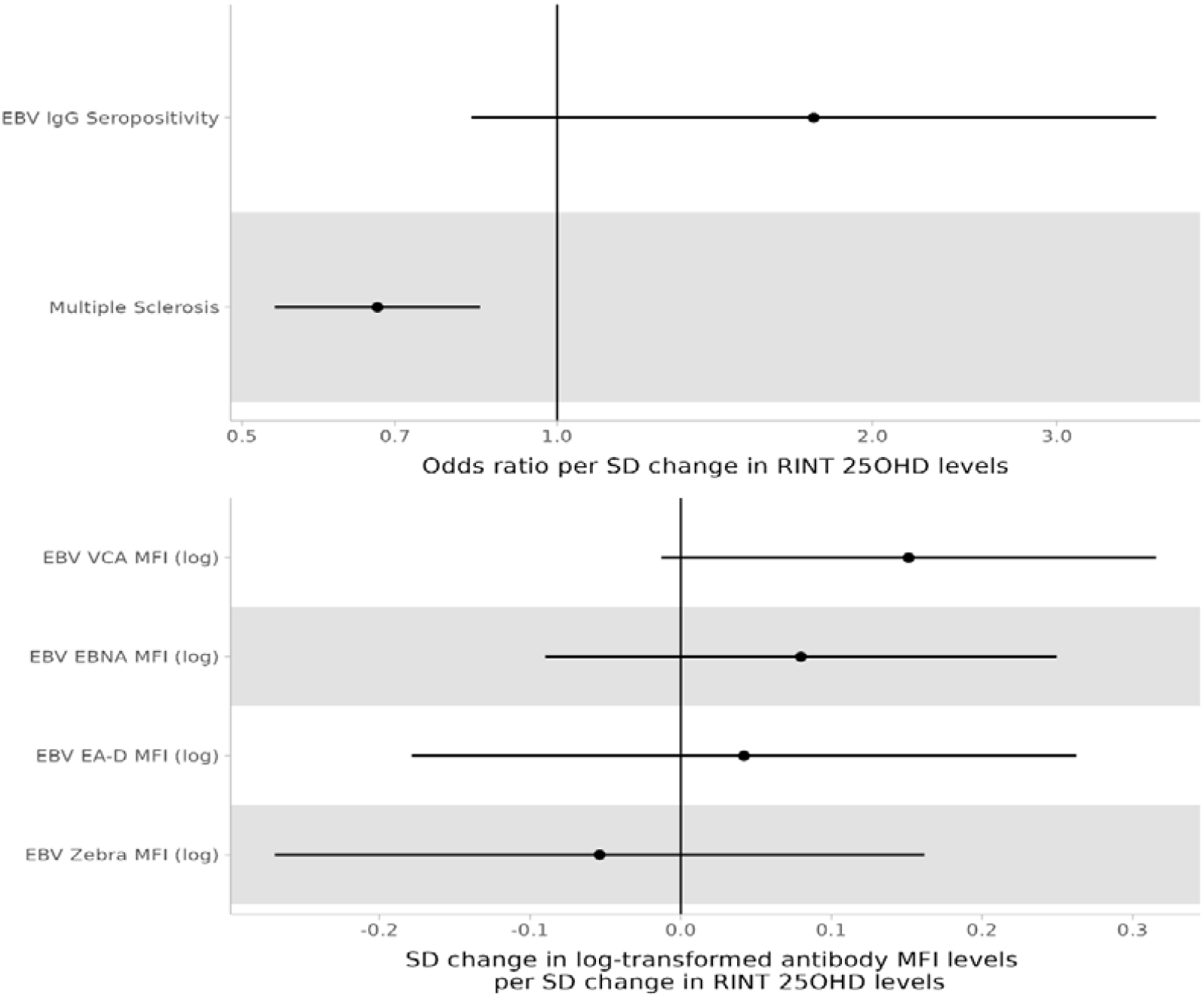
Mendelian Randomization of vitamin D on serological evidence of EBV and Multiple Sclerosis.

With the exception of ZEBRA, for which there was a -0.05-fold SD change in log transformed antibody MFI per SD change in RINT vitamin D levels (95% CI -0.27 to 0.16), all EBV serological had positive MR estimates for genetically proxied vitamin D levels (beta ranging from 0.04 (95% CI -0.18 to 0.26) for EA-D to 0.15 (95% CI - 0.01 to 0.32) for VCA. In addition, IgG seropositivity also had a positive MR estimate, with an OR of 1.76 per SD increase in genetically proxied vitamin D (95% CI 0.83-3.73). In all instances however the 95% CI crossed the null value.

When we repeated the analysis without restricting to SNPs *cis* to genes known to be involved in vitamin D synthesis the interpretation of the effect of genetically proxied vitamin D levels on serological markers of EBV infection was unchanged, with the 95% CI for all estimates crossing the null (Supplementary Figure 1). Similar magnitudes and directions of effect were noted when using other MR analyses (Supplementary Figure 2). There was evidence of heterogeneity for the IVs for the analysis of EBV EA-D (Cochran’s Q = 27.09, p 0.04) and ZEBRA antibody levels (Cochran’s Q = 27.24, p 0.04) (Supplementary Table 4). For all other serological markers and MS case status there was no detectable heterogeneity. MR-Egger intercept tests provided no evidence of directional horizontal pleiotropy for any EBV serology analysis or for the MS case status analysis (Supplementary Table 5).

**Figure 2.**
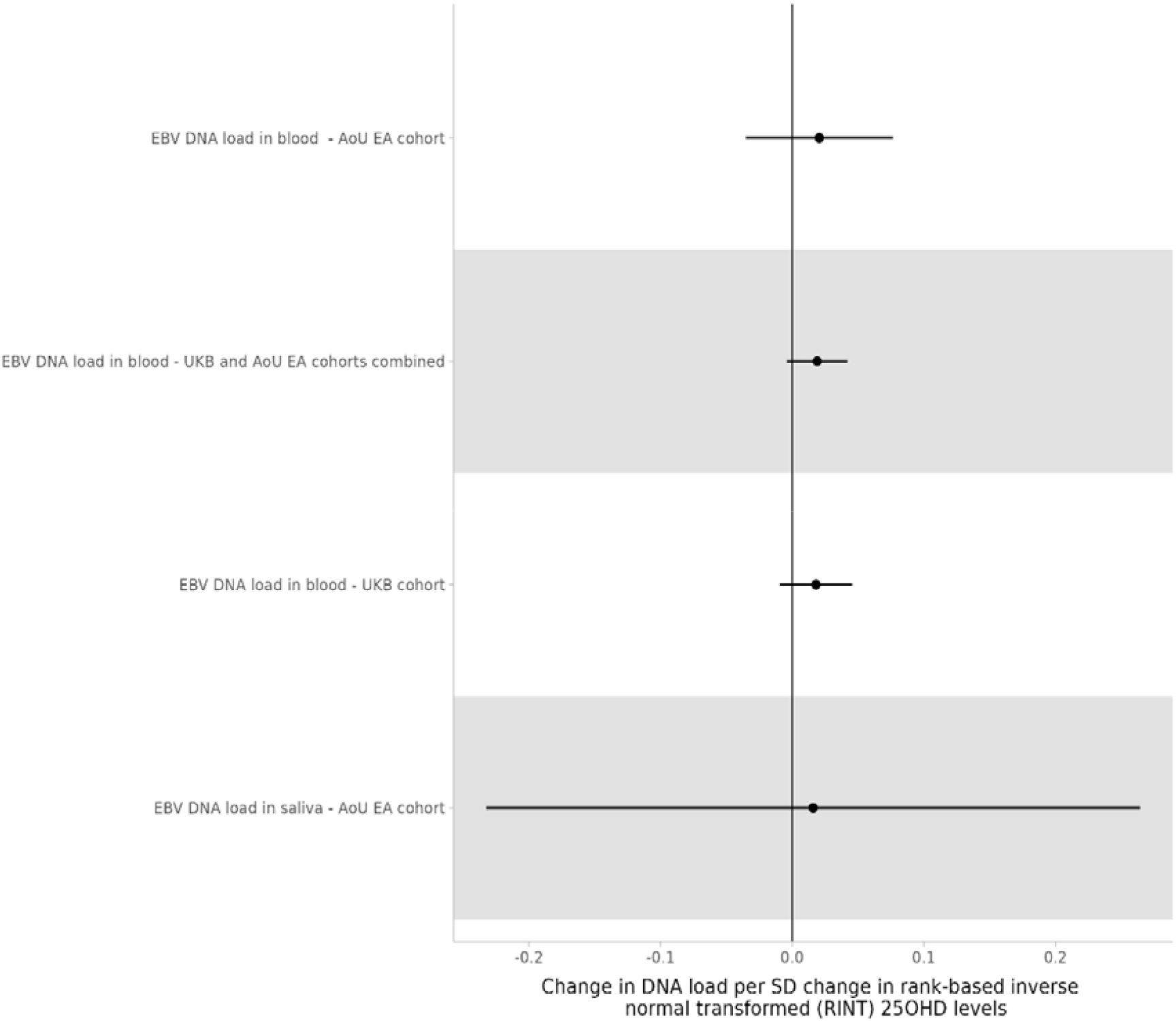
Mendelian Randomization of vitamin D on EBV DNA load in blood and saliva.

### Detectable EBV DNA

There was no evidence of an increase in detectable EBV DNA in both saliva and blood with higher genetically proxied vitamin D levels (Figure 2).

For the combined cohort of European Ancestry (EA) individuals from both UKB and AoU an SD increase in RINT transformed vitamin D levels caused a 0.02-fold increase in the INT count of EBV DNA read pairs (95% CI -0.004 to 0.04) with a CI that crossed the null. This combined estimate is in line with the individual estimates UKB (beta 0.02, 95% CI (-0.009 to 0.05) and AoU EA cohorts (beta 0.02 (95%

CI -0.03 to 0.08).

For detectable EBV DNA in saliva the effect estimate was similar to that for blood, with a small estimated effect and CI that crossed the null (0.02-fold increase in INT count of EBV DNA read pairs (95% CI -0.23 to 0.26) per SD increase in RINT 25OH vitamin D.

When the analysis was not restricted to SNPs *cis* to genes known to be involved in vitamin D synthesis the findings were similar (Supplementary Figure 3). When comparing different MR approaches the direction and magnitude of effect for detectable EBV DNA in blood in the combined EA cohort was similar across all (Supplementary Figure 4). There was no evidence of heterogeneity amongst the IVs across all MR IVW analyses of EBV DNA load (Supplementary Table 4). MR-Egger intercept tests provided no evidence of directional horizontal pleiotropy for any EBV DNA load analysis (Supplementary Table 5).

## Discussion

We found no definitive evidence that genetically proxied vitamin D levels are causally linked to the likelihood of either serological evidence of EBV infection or detectable EBV DNA in peripheral blood or saliva. We confirm the finding, documented extensively elsewhere, that lower vitamin D levels are causally linked to risk of developing MS.

EBV seropositivity increases with latitude at the population level(35). Furthermore, EBV DNAemia increases in individuals born in more Northern latitudes(25). We fail to show an effect of vitamin D levels on either serological evidence of EBV infection or EBV DNAemia, with our direction of effect in fact suggesting higher EBV DNA load in blood with higher vitamin D, although this effect size was small and the CI crossed the null. This might mean that effect of latitude on EBV infection prevalence is not entirely explained by vitamin D levels. Alternatively it might reflect our lack of data regarding the age of EBV acquisition, which is likely a key factor in the development of MS(16).

The initial work on EBV DNAemia showed differing relationships between detectable EBV DNA in blood and serological markers of EBV infection, with strong concordance with quantitative antibody responses to VCA-p18 and ZEBRA, weak effects to EA-D and no effect to EBNA-1, illustrating differing immunological responses to viral DNA load(26). This work also showed that EBV DNA viral load in blood was positively correlated with development of Hodgkin’s lymphoma, but not MS(26), highlighting that there are likely various, disease-specific mechanisms linking EBV infection and development of associated conditions. Given that EBV DNA viral load in blood was not found to be associated with MS in this study, it is perhaps unsurprising that we fail to show a causal effect of vitamin D upon EBV DNAemia.

Given that we confirm the effect of vitamin D on development of MS, alongside the well documented association between EBV and MS documented elsewhere our failure to demonstrate a link between vitamin D and EBV may suggest that vitamin D and EBV act via separate pathways on the risk of MS. Alternatively there could be a link which is mediated through an uninvestigated pathway. Autoantibodies which exhibit cross-reactivity between EBNA1 (Epstein-Barr nuclear antigen 1) and self-antigens including myelin basic protein demonstrate how EBV could hypothetically trigger MS(36). We did not assess the presence of autoantibodies. Previous work suggested that vitamin D supplementation in individuals with MS reduced levels of EBNA-1 antibody levels but not VCA(37) so it is interesting that we failed to show an effect for either, although our work was not restricted to only those with MS.

This is the first published MR analysis to our knowledge to investigate the link between vitamin D levels and EBV infection, using large, well-powered datasets. The use of EBV DNAemia is a strength, providing an additional dimension to conventional serological quantification of infection. In addition the MR design reduces the likelihood of residual confounding, exposure misclassification and reverse causation which other designs might encounter. Finally we restrict our analysis to SNPs that are *cis* to genes known to directly influence vitamin D levels, reducing the likelihood of horizontal pleiotropy.

There are limitations to this work. Firstly infection is a dynamic process, evolving over time and in the context of the host response to infection(38). Antibody levels capture historical evidence of infection but are inexact, with false positives possible due to cross reactivity with other antibodies and false negatives possible in individuals with waning antibody levels, or who failed to ever produce antibodies, for instance due to immunosuppression. This could theoretically bias towards a null result, as we present.

Measurement of EBV DNA goes some way towards addressing this limitation, however again only reflects a single timepoint. EBV DNAemia exhibits seasonal and circadian variation, with intermittent detectability at the level of the individual(24, 26). Further, factors associated with EBV DNAemia, including current smoking and the use of immunosuppressive drugs(39) can vary over time, potentially limiting the extent to which a single assessment accurately captures control of EBV DNAemia. Furthermore detectability is dependent upon technical factors. These include the volume of extracted DNA and depth of sequencing. Therefore, there is potential measurement error in the use of EBV DNAemia as an outcome.

Secondly EBV is common in the adult population with incidence increasing with age. It typically affects individuals at a young age, with approximately 60% of 11-14 year olds infected(40). The median age of those analysed in our serological outcome dataset was 58, with consequent high seroprevalence (94.4%) and no data on age of acquisition(19). Infection with EBV during a vulnerable period in late adolescence or early adulthood is a key variable in determining the development of MS(16, 22), yet we lack data on age of infection here and age of initiation of MS. Vitamin D levels have been shown to associate with reactivity against EBNA-1 (which is itself associated with MS risk) most strongly in those below 26.4 years of age. Vitamin D levels in early life may be the key exposure variable(41), potentially in the subset of those exposed to EBV during the vulnerable period.

Our IVs are taken from a population aged between 40-69 years and we assume that our instruments affect vitamin D levels across the life course. However we lack information on vitamin D levels in early life to confirm this and there is no current life-course GWAS of vitamin D. Life course GWAS applied to the effect of adiposity between menarche and first pregnancy on breast cancer risk, demonstrated heterogeneity in effects with distinct genetic effects at specific time points(42). It is possible that genetic effects mediate vitamin D differentially between childhood and adulthood, which our study would miss.

Finally our instruments were derived from UKB, as were many of our outcomes, meaning there will be strong sample overlap. This could bias the results to the observational. It should be noted here that we present a null effect, so bias towards the observational association is unlikely to explain the absence of an effect.

This work provides no evidence to support a causal link between vitamin D levels and infection status with EBV. Future work could measure EBV DNA and vitamin D levels at multiple timepoints across the vulnerable period between childhood and early adulthood alongside MS case status, which would theoretically be possible from large cohorts with stored samples or from a de novo trial.

## Supporting information

MR-STROBE Form

## Data Availability

All data produced are available online as described in the supplementary material

## Data and data sharing

The Vitamin D GWAS is available at https://www.ebi.ac.uk/gwas/studies/GCST9000061. EBV serology was obtained via the following links: IgG seropositivity (ebi-a-GCST90006897 - Anti-Epstein-Barr virus IgG seropositivity - OpenGWAS), VCA p18 antibody level (ebi-a-GCST90006900 - Epstein-Barr virus VCA p18 antibody levels - OpenGWAS), ZEBRA antibody (ebi-a-GCST90006901 - Epstein-Barr virus ZEBRA antibody levels - OpenGWAS), EBNA- 1 antibody levels (ebi-a-GCST90006899 - Epstein-Barr virus EBNA-1 antibody levels - OpenGWAS), EA-D antibody levels (ebi-a-GCST90006898 - Epstein-Barr virus EA-D antibody levels - OpenGWAS). Multiple sclerosis case status is available at ieu-b-18 - multiple sclerosis - OpenGWAS.

Detectable EBV DNA in blood or saliva is from GCST90809801-7 available at https://www.ebi.ac.uk/gwas/studies/GCST90809801

Statistical code will be uploaded to the lead author’s GitHub

