## Supplementary material for "Serum Vitamin D Levels do not Affect Susceptibility to EBV Infection – A Mendelian Randomization Study": MR-STROBE Form

**STROBE-MR checklist of recommended items to address in reports of Mendelian randomization studies**^1^ ^2^

| **Item No.** | **Section** | **Checklist item** | **Page No.** | **Relevant text from manuscript** |
| --- | --- | --- | --- | --- |
| 1 | **TITLE and ABSTRACT** | Indicate Mendelian randomization (MR) as the study’s design in the title and/or the abstract if that is a main purpose of the study | 1 | Title – "Serum Vitamin D Levels do not Affect Susceptibility to EBV Infection – A Mendelian Randomization Study." |
|  | **INTRODUCTION** |  |  |  |
| 2 | **Background** | Explain the scientific background and rationale for the reported study. What is the exposure? Is a potential causal relationship between exposure and outcome plausible? Justify why MR is a helpful method to address the study question | 2-3 | Extensive discussion of the issues of reverse causality and residual confounding in the vitamin D literature. |
| 3 | **Objectives** | State specific objectives clearly, including pre-specified causal hypotheses (if any). State that MR is a method that, under specific assumptions, intends to estimate causal effects | 2 | "We hypothesize that vitamin D levels may influence immune control of EBV and explain the as-yet unexplained association between vitamin D and MS."  "We aim to investigate, using mendelian randomization (MR)(26, 27), whether serum vitamin D levels are causally linked to a) serological evidence of EBV infection and b) detectable EBV DNA in blood and saliva." |
|  | **METHODS** |  |  |  |
| 4 | **Study design and data sources** | Present key elements of the study design early in the article. Consider including a table listing sources of data for all phases of the study. For each data source contributing to the analysis, describe the following: | 15/16 | Table 1 |
|  | a) | Setting: Describe the study design and the underlying population, if possible. Describe the setting, locations, and relevant dates, including periods of recruitment, exposure, follow-up, and data collection, when available. | 15/16 | Table 1 |
|  | b) | Participants: Give the eligibility criteria, and the sources and methods of selection of participants. Report the sample size, and whether any power or sample size calculations were carried out prior to the main analysis | 15/16 | Table 1 |
|  | c) | Describe measurement, quality control and selection of genetic variants | 6 | "We restricted instruments to those that are cis (within 10^6^ base pairs) to genes known to be on the vitamin D pathway (*CYP2R1, GC, NADSYN1/ DHCR7, CYP24A1*), independent (R^2^ > 0.1) and strongly associated with vitamin D blood levels (p < 5 x 10^-8^)." |
|  | d) | For each exposure, outcome, and other relevant variables, describe methods of assessment and diagnostic criteria for diseases | 6 | "In this GWAS individuals had total 25OHD (combined 25OHD_3_ and 25OHD_2_) measured using the Diasorin Liason®, a chemiluminescent immunoassay (CLIA)." |
|  | e) | Provide details of ethics committee approval and participant informed consent, if relevant | 2 | " Ethics: No formal ethics approval was needed for this study as publicly available data was used." |
| 5 | **Assumptions** | Explicitly state the three core IV assumptions for the main analysis (relevance, independence and exclusion restriction) as well assumptions for any additional or sensitivity analysis | 6 | To satisfy the three core instrumental variable (IV) assumptions for MR in an IV framework(26) of relevance, independence and exclusion restriction we applied the following criteria to our IVs. We restricted instruments to those that are cis (within 10^6^ base pairs) to genes known to be on the vitamin D pathway (*CYP2R1, GC, NADSYN1/ DHCR7, CYP24A1*), independent (R^2^ > 0.1) and strongly associated with vitamin D blood levels (p < 5 x 10^-8^). |
| 6 | **Statistical methods: main analysis** | Describe statistical methods and statistics used |  |  |
|  | a) | Describe how quantitative variables were handled in the analyses (i.e., scale, units, model) | 5 | Methods and Table 1 |
|  | b) | Describe how genetic variants were handled in the analyses and, if applicable, how their weights were selected | 6 | " We restricted instruments to those that are cis (within 10^6^ base pairs) to genes known to be on the vitamin D pathway (*CYP2R1, GC, NADSYN1/ DHCR7, CYP24A1*), independent (R^2^ > 0.1) and strongly associated with vitamin D blood levels (p < 5 x 10^-8^).”  Weights = NA |
|  | c) | Describe the MR estimator (e.g. two-stage least squares, Wald ratio) and related statistics. Detail the included covariates and, in case of two-sample MR, whether the same covariate set was used for adjustment in the two samples | 8 | "Estimates were generated by the Wald Ratio and analysed by inverse-variance weighting." |
|  | d) | Explain how missing data were addressed | 7 | Exposure-associated SNPs that were missing in the outcome dataset were excluded from the analysis |
|  | e) | If applicable, indicate how multiple testing was addressed | - | NA |
| 7 | **Assessment of assumptions** | Describe any methods or prior knowledge used to assess the assumptions or justify their validity | 5  11 | Relevance – ". We restricted instruments to those that are cis (within 10^6^ base pairs) to genes known to be on the vitamin D pathway (*CYP2R1, GC, NADSYN1/ DHCR7, CYP24A1*), independent (R^2^ > 0.1) and strongly associated with vitamin D blood levels (p < 5 x 10^-8^)"  Exclusion restriction – ". MR-Egger intercept tests provided no evidence of directional horizontal pleiotropy for any detectable EBV DNA analysis (Supplementary Table 2)" |
| 8 | **Sensitivity analyses and additional analyses** | Describe any sensitivity analyses or additional analyses performed (e.g. comparison of effect estimates from different approaches, independent replication, bias analytic techniques, validation of instruments, simulations) | 6 | "We repeated our IVW MR with IVs as described above, including the requirements for independence and strong association with vitamin D levels but omitting the requirement for the SNPs to be cis to genes known to be on the Vitamin D pathway.  We also report the results of MR using Egger, Simple Mode, Weighted Mode and Weighted Median as sensitivity analyses. To assess for pleiotropy we performed MR Egger and report the p value and intercept. To assess for heterogeneity we report Cochran's Q for the IVW analysis." |
| 9 | **Software and pre-registration** |  |  |  |
|  | a) | Name statistical software and package(s), including version and settings used | 7 | The analysis was performed in R version 4.5.3(30) using the Two-Sample MR package(31). |
|  | b) | State whether the study protocol and details were pre-registered (as well as when and where) | 7 | "This study was not pre-registered." |
|  | **RESULTS** |  |  |  |
| 10 | **Descriptive data** |  |  |  |
|  | a) | Report the numbers of individuals at each stage of included studies and reasons for exclusion. Consider use of a flow diagram | 8 | After restriction as detailed above we had 17 SNPs as IVs. These corresponded to the following numbers of SNPs for each of our outcomes:   - 17 SNPs for all EBV serological markers - 14 SNPS for MS case status - 8 SNPs for all EBV in blood/saliva analyses, with the exception of the African ancestry EBV DNA load in the blood, for which there were 4 SNPs. |
|  | b) | Report summary statistics for phenotypic exposure(s), outcome(s), and other relevant variables (e.g. means, SDs, proportions) | 15/16 | Table 1 |
|  | c) | If the data sources include meta-analyses of previous studies, provide the assessments of heterogeneity across these studies | - | NA |
|  | d) | For two-sample MR:  i.  Provide justification of the similarity of the genetic variant-exposure associations between the exposure and outcome samples  ii.  Provide information on the number of individuals who overlap between the exposure and outcome studies |  | (i) Both exposure and outcome are from similarly aged European ancestry individuals  ii) Overlap not known but mentioned in discussion - " our instruments were derived from UKB, as were many of our outcomes, meaning there will be strong sample overlap. This could bias the results to the observational. It should be noted here that we present a null effect." |
| 11 | **Main results** |  |  |  |
|  | a) | Report the associations between genetic variant and exposure, and between genetic variant and outcome, preferably on an interpretable scale |  | Supplementary Tables 1 and 2 |
|  | b) | Report MR estimates of the relationship between exposure and outcome, and the measures of uncertainty from the MR analysis, on an interpretable scale, such as odds ratio or relative risk per SD difference |  | " the measured levels (log median fluorescence intensity (MFI)) of all EBV serological markers increased with higher genetically proxied vitamin D levels (OR ranging from 1.04 (95% CI 0.84-1.30) for EA-D to 1.16 (95% CI 0.99 – 1.37) for VCA p18 per SD change in RINT-transformed vitamin D levels)" |
|  | c) | If relevant, consider translating estimates of relative risk into absolute risk for a meaningful time period | - | NA |
|  | d) | Consider plots to visualize results (e.g. forest plot, scatterplot of associations between genetic variants and outcome versus between genetic variants and exposure) | 9 & 12 | Figures 1 and 2 |
| 12 | **Assessment of assumptions** |  |  |  |
|  | a) | Report the assessment of the validity of the assumptions |  |  |
|  | b) | Report any additional statistics (e.g., assessments of heterogeneity across genetic variants, such as *I^2^*, Q statistic or E-value) | 11 | " There was no evidence of heterogeneity amongst the IVs across all MR IVW analyses of EBV DNA load (Supplementary Table 1). MR-Egger intercept tests provided no evidence of directional horizontal pleiotropy for any detectable EBV DNA analysis (Supplementary Table 2). For the EBV 1 versus 2 analysis there was suggestion of directional horizontal pleiotropy (intercept = 0.013, p = 0.026)." |
| 13 | **Sensitivity analyses and additional analyses** |  |  |  |
|  | a) | Report any sensitivity analyses to assess the robustness of the main results to violations of the assumptions |  | Supplementary Figures 1 and 3 |
|  | b) | Report results from other sensitivity analyses or additional analyses |  | Supplementary Figures 2 and 4 |
|  | c) | Report any assessment of direction of causal relationship (e.g., bidirectional MR) | - | NA |
|  | d) | When relevant, report and compare with estimates from non-MR analyses | - | NA |
|  | e) | Consider additional plots to visualize results (e.g., leave-one-out analyses) |  |  |
|  | **DISCUSSION** |  |  |  |
| 14 | **Key results** | Summarize key results with reference to study objectives | 11 | "We found no definitive evidence that genetically proxied vitamin D levels are causally linked to the likelihood of either serological evidence of EBV infection or detectable EBV DNA in peripheral blood or saliva" |
| 15 | **Limitations** | Discuss limitations of the study, taking into account the validity of the IV assumptions, other sources of potential bias, and imprecision. Discuss both direction and magnitude of any potential bias and any efforts to address them |  | Extensive discussion of limitations |
| 16 | **Interpretation** |  |  |  |
|  | a) | Meaning: Give a cautious overall interpretation of results in the context of their limitations and in comparison with other studies | 10 | "The initial work on EBV DNAemia …… showed that EBV DNA viral load in blood was positively correlated with development of Hodgkin's lymphoma, but not MS(24), highlighting that there are likely various, disease-specific mechanisms linking EBV infection and development of associated conditions. Given that EBV DNA viral load in blood was not found to be associated with MS in this study, it is perhaps unsurprising that we fail to show a causal effect of vitamin D upon EBV DNAemia." |
|  | b) | Mechanism: Discuss underlying biological mechanisms that could drive a potential causal relationship between the investigated exposure and the outcome, and whether the gene-environment equivalence assumption is reasonable. Use causal language carefully, clarifying that IV estimates may provide causal effects only under certain assumptions | 13 | "(23). We fail to show an effect of vitamin D levels on either serological evidence of EBV infection or EBV DNAemia. This might mean that effect of latitude on EBV infection prevalence is not entirely explained by vitamin D levels. Alternatively it might reflect our lack of data regarding the age of EBV acquisition, which is likely a key factor in the development of MS(15). Indeed our vitamin D IVs are taken from a population aged between 40-69 years, much later than the vulnerable period of adolescence and early adulthood during which EBV acquisition predisposes to the development of MS. It may be that vitamin D levels in early life, for which we lack the data, are causally linked to EBV infection." |
|  | c) | Clinical relevance: Discuss whether the results have clinical or public policy relevance, and to what extent they inform effect sizes of possible interventions | 13 | "Future work could measure EBV DNA and vitamin D levels at multiple timepoints across the vulnerable period between childhood and early adulthood alongside MS case status" |
| 17 | **Generalizability** | Discuss the generalizability of the study results (a) to other populations, (b) across other exposure periods/timings, and (c) across other levels of exposure |  | A) + B)" our vitamin D IVs are taken from a population aged between 40-69 years, much later than the vulnerable period of adolescence and early adulthood during which EBV acquisition predisposes to the development of MS. It may be that vitamin D levels in early life, for which we lack the data, are causally linked to EBV infection." |
|  | **OTHER INFORMATION** |  |  |  |
| 18 | **Funding** | Describe sources of funding and the role of funders in the present study and, if applicable, sources of funding for the databases and original study or studies on which the present study is based | 2 | " Funding: This study was funded by the NIHR Clinical Lectureship programme (FH) and the NIHR Academic Clinical Fellowship programme (JS). The research was carried out at the National Institute for Health and Care Research (NIHR) Bristol Biomedical Research Centre (BRC). The funders had no role in inception, analysis or reporting of the study." |
| 19 | **Data and data sharing** | Provide the data used to perform all analyses or report where and how the data can be accessed, and reference these sources in the article. Provide the statistical code needed to reproduce the results in the article, or report whether the code is publicly accessible and if so, where | 12 | Data sources provided |
| 20 | **Conflicts of Interest** | All authors should declare all potential conflicts of interest | 2 | "Conflict of Interest: No authors have any conflict of interest to report." |

This checklist is copyrighted by the Equator Network under the Creative Commons Attribution 3.0 Unported (CC BY 3.0) license.

1.	Skrivankova VW, Richmond RC, Woolf BAR, Yarmolinsky J, Davies NM, Swanson SA, et al. Strengthening the Reporting of Observational Studies in Epidemiology using Mendelian Randomization (STROBE-MR) Statement. JAMA. 2021;under review.

2.	Skrivankova VW, Richmond RC, Woolf BAR, Davies NM, Swanson SA, VanderWeele TJ, et al. Strengthening the Reporting of Observational Studies in Epidemiology using Mendelian Randomisation (STROBE-MR): Explanation and Elaboration. BMJ. 2021;375:n2233.
