## Supplementary Information for "Serum Vitamin D Levels do not Affect Susceptibility to EBV Infection – A Mendelian Randomization Study"

**Supplementary Material**

**Supplementary Figure 1. Mendelian Randomization of vitamin D (all SNPs) on serological evidence of EBV and Multiple Sclerosis**

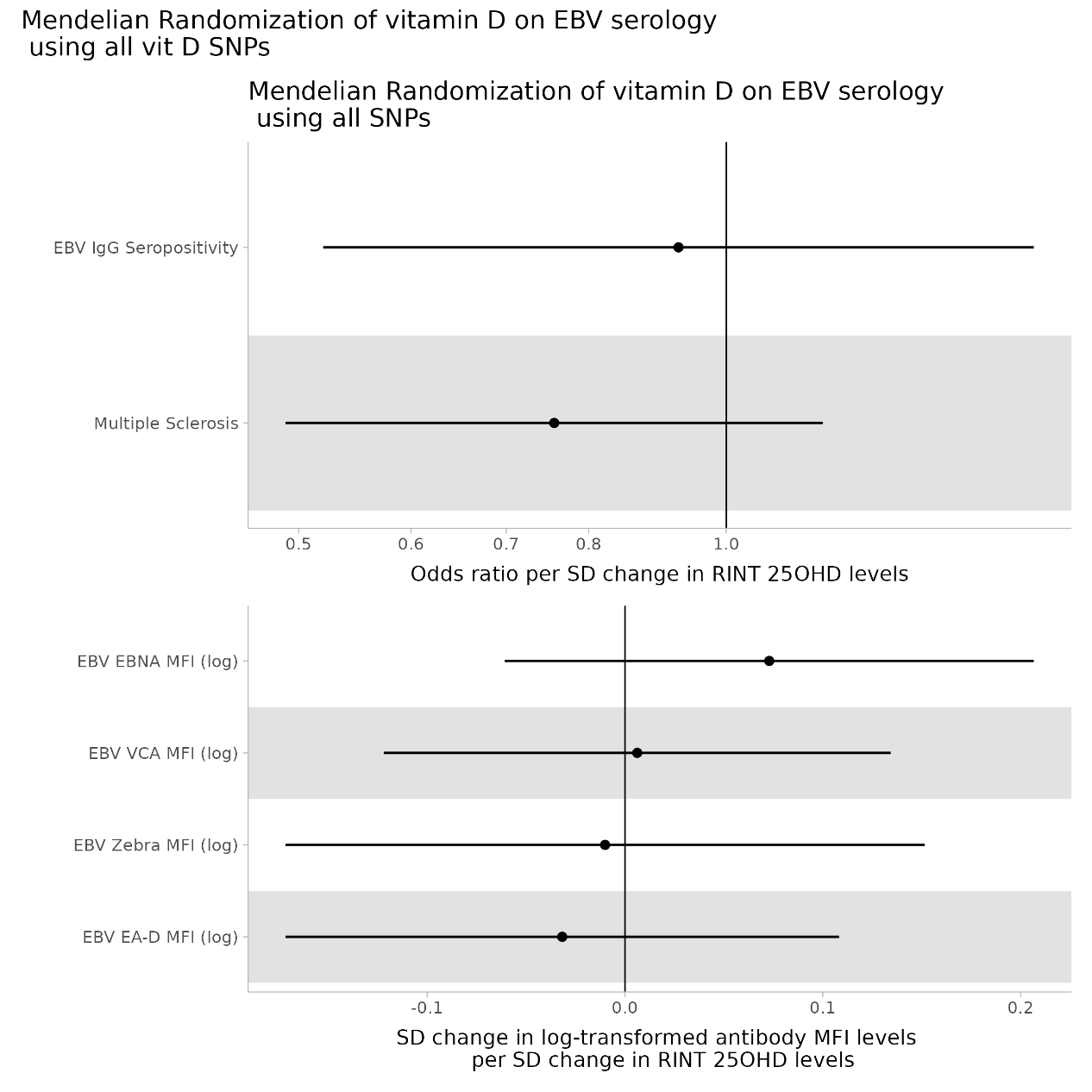

**Supplementary Figure 2. Scatter Plots of Mendelian Randomization of vitamin D on EBV serological markers and MS.**

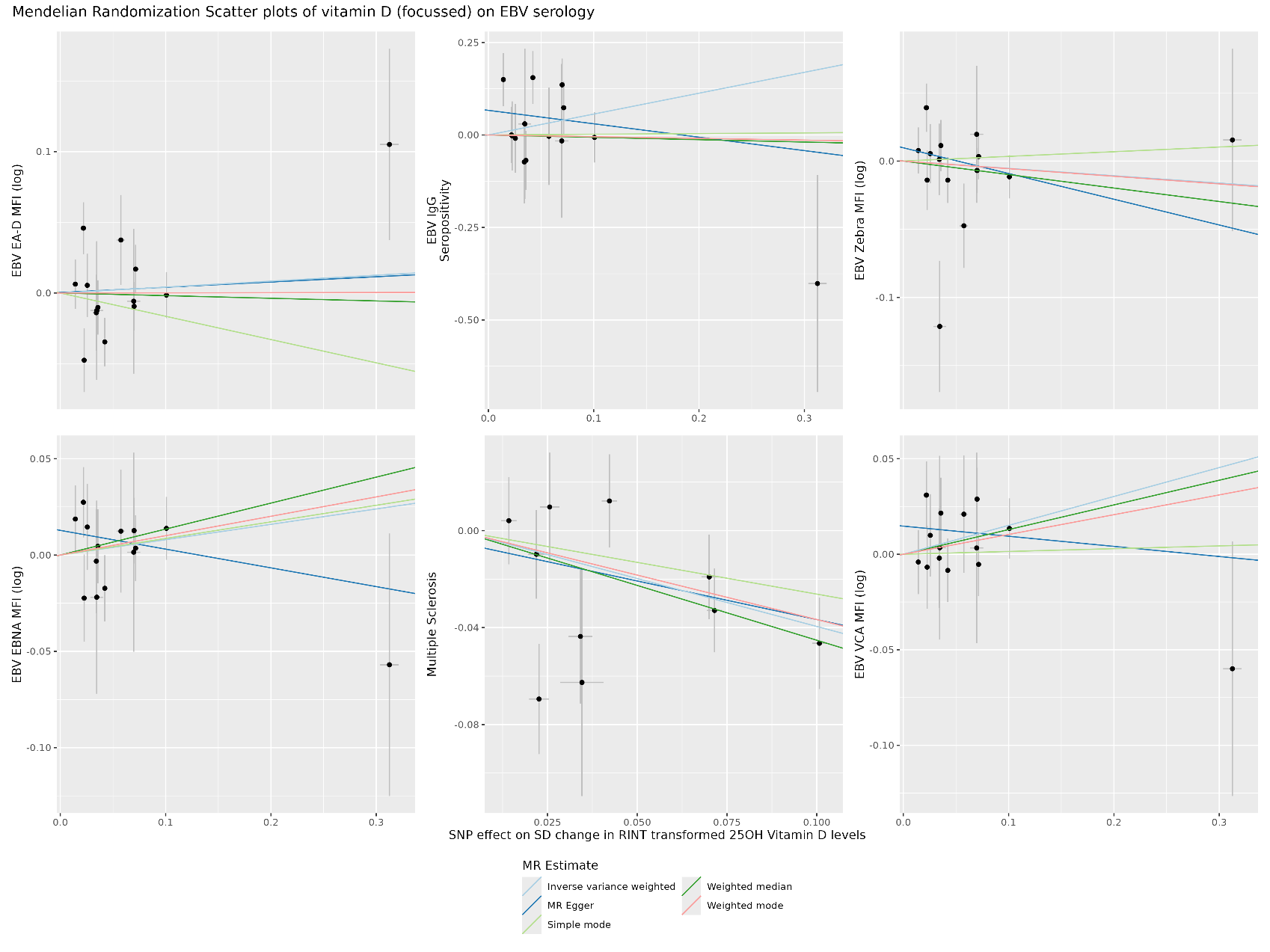

**Supplementary Figure 3. Mendelian Randomization of vitamin D (all SNPS) on detectable EBV in blood and saliva and EBV serotype.**

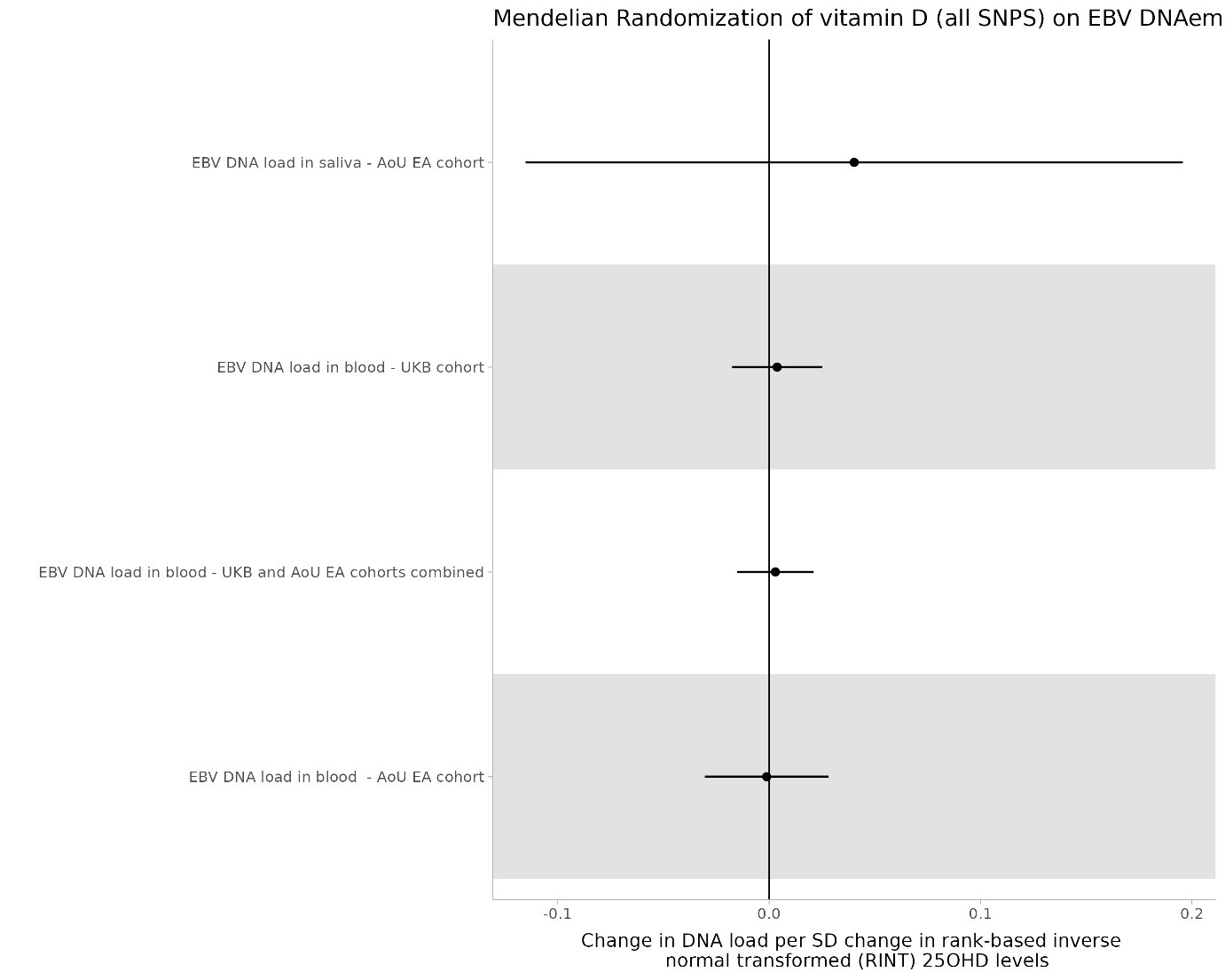

**Supplementary Figure 4. Scatter Plots of Mendelian Randomization of vitamin D on detectable EBV in blood and saliva and EBV serotype.**

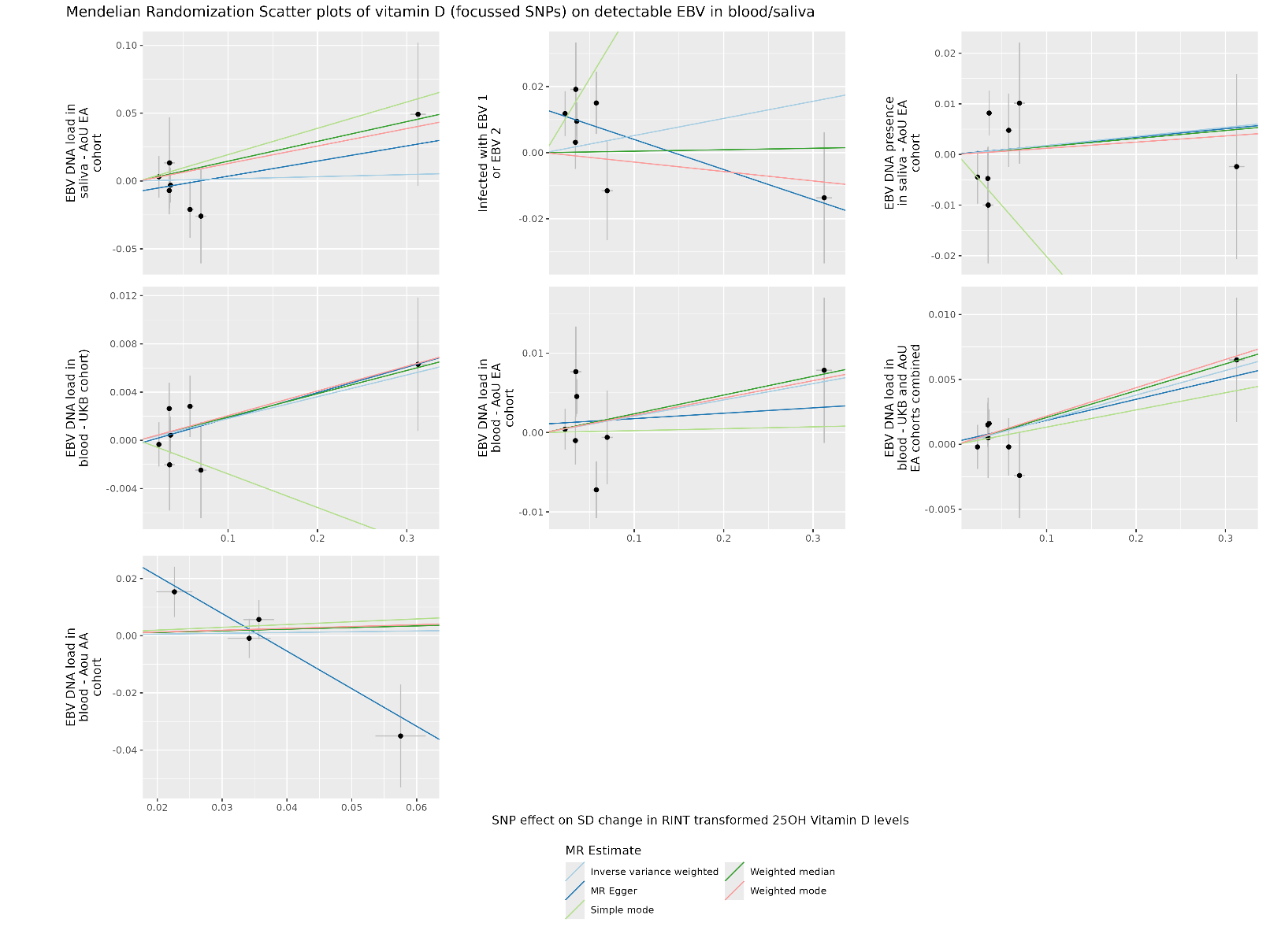

| **Supplementary Table 1. Sources of data for MR analysis** | | | |  |  |
| --- | --- | --- | --- | --- | --- |
| **Paper** | **GWAS ID** | **Trait and units** | **Age** | **Dataset** | **N** |
| Revez et al | ebi-a-[GCST90000616](https://www.ebi.ac.uk/gwas/studies/GCST90000616) | Total 25OHD Vitamin D – rank-based inverse-normal transformation (RINT transformed) | Range of 40-69 | UK Biobank – enrolled 2006-10 | 417,580 European ancestry individuals |
| Butler-Laporte 2020 | [ebi-a-GCST90006897](https://gwas.mrcieu.ac.uk/datasets/ebi-a-GCST90006897/) | IgG seropositivity (positive for 2 or more antigens) | Median 58, interquartile range 51-64 | UK Biobank – enrolled 2006-10 | 8477 European ancestry individuals |
|  | [ebi-a-GCST90006900](https://gwas.mrcieu.ac.uk/datasets/ebi-a-GCST90006900/) | VCA p18 antibody level – units = standardised median fluorescence intensity (MFI) (log)~ |  |  | 8518 European ancestry individuals |
|  | [ebi-a-GCST90006901](https://gwas.mrcieu.ac.uk/datasets/ebi-a-GCST90006901/) | ZEBRA antibody level – MFI(log) |  |  | 8191 European ancestry individuals |
|  | [ebi-a-GCST90006899](https://gwas.mrcieu.ac.uk/datasets/ebi-a-GCST90006899/) | EBNA-1 antibody level – MFI(log) |  |  | 7972 European ancestry individuals |
|  | [ebi-a-GCST90006898](https://gwas.mrcieu.ac.uk/datasets/ebi-a-GCST90006898/) | EA-D antibody level – log MFI(log) |  |  | 7763 European ancestry individuals |
| Patsopoulos 2019 | ieu-b-18 | Multiple Sclerosis case status | Varied by cohort. Typically over the age of 16 | International Multiple Sclerosis Genetics Consortium GWAS and national MS cohorts | 47,429 multiple sclerosis (MS) cases and 68,374 controls from combining 15 GWAS across Australia, US and Europe. |
| Kamitaki 2026 | ebi-a-GCST90809806 | EBV DNA load in saliva - AoU EA cohort* | Mean 53.1, standard deviation (SD) 17(32) | All of Us – enrolment of US individuals from 2018 onwards | 16,282 European ancestry individuals |
|  | ebi-a-GCST90809802 | EBV DNA load in blood - AoU EA cohort* |  |  | 201,168 European ancestry individuals (US) |
|  | ebi-a-GCST90809803 | EBV DNA load in blood – UKB and AoU EA cohorts combined* | Not available but each database summarised above/below | Combined UKB and AoU European cohorts | 654,938 European ancestry individuals (US, UK) |
|  | ebi-a-GCST90809804 | EBV DNA load in blood – UKB cohort* | Range of 40 - 69 | UK Biobank – enrolled 2006-10 | 453,770 European ancestry individuals (UK) |
| (~) For serological markers the log MFI was standardised to give mean of 0 and SD of 1  (*) For all viral DNA load outcomes the units are inverse normal transformed count of read pairs. | | | | | |

| **Supplementary Table 2 - SNP level effect estimates for MR of vitamin D on EBV serological markers and MS** | | | | | | | |
| --- | --- | --- | --- | --- | --- | --- | --- |
| SNP | outcome | beta.exposure | se.exposure | pval.exposure | beta.outcome | se.outcome | pval.outcome |
| rs11023212 | EBV EA-D MFI (log) | 0.072 | 0.002 | <5e-08 | 0.017 | 0.017 | 0.325 |
| rs117592720 | EBV EA-D MFI (log) | -0.070 | 0.006 | <5e-08 | 0.006 | 0.051 | 0.910 |
| rs146128209 | EBV EA-D MFI (log) | 0.058 | 0.004 | <5e-08 | 0.037 | 0.032 | 0.239 |
| rs1792287 | EBV EA-D MFI (log) | -0.022 | 0.002 | <5e-08 | -0.046 | 0.018 | 0.012 |
| rs2585442 | EBV EA-D MFI (log) | -0.036 | 0.002 | <5e-08 | 0.010 | 0.019 | 0.596 |
| rs2616279 | EBV EA-D MFI (log) | 0.023 | 0.003 | <5e-08 | -0.048 | 0.023 | 0.035 |
| rs4694423 | EBV EA-D MFI (log) | 0.101 | 0.002 | <5e-08 | -0.002 | 0.016 | 0.926 |
| rs4944885 | EBV EA-D MFI (log) | -0.070 | 0.002 | <5e-08 | 0.009 | 0.017 | 0.579 |
| rs543746207 | EBV EA-D MFI (log) | 0.312 | 0.009 | <5e-08 | 0.105 | 0.068 | 0.121 |
| rs6123359 | EBV EA-D MFI (log) | -0.034 | 0.003 | <5e-08 | 0.014 | 0.027 | 0.604 |
| rs71467497 | EBV EA-D MFI (log) | 0.035 | 0.006 | <5e-08 | -0.012 | 0.049 | 0.800 |
| rs71601787 | EBV EA-D MFI (log) | -0.042 | 0.002 | <5e-08 | 0.035 | 0.017 | 0.044 |
| rs7657132 | EBV EA-D MFI (log) | 0.014 | 0.002 | <5e-08 | 0.006 | 0.017 | 0.719 |
| rs7675923 | EBV EA-D MFI (log) | -0.026 | 0.003 | <5e-08 | -0.005 | 0.022 | 0.808 |
| rs11023212 | EBV EBNA MFI (log) | 0.072 | 0.002 | <5e-08 | 0.004 | 0.017 | 0.836 |
| rs117592720 | EBV EBNA MFI (log) | -0.070 | 0.006 | <5e-08 | -0.001 | 0.052 | 0.977 |
| rs146128209 | EBV EBNA MFI (log) | 0.058 | 0.004 | <5e-08 | 0.012 | 0.032 | 0.698 |
| rs1792287 | EBV EBNA MFI (log) | -0.022 | 0.002 | <5e-08 | -0.027 | 0.018 | 0.133 |
| rs2585442 | EBV EBNA MFI (log) | -0.036 | 0.002 | <5e-08 | -0.005 | 0.019 | 0.813 |
| rs2616279 | EBV EBNA MFI (log) | 0.023 | 0.003 | <5e-08 | -0.022 | 0.023 | 0.324 |
| rs4694423 | EBV EBNA MFI (log) | 0.101 | 0.002 | <5e-08 | 0.014 | 0.016 | 0.395 |
| rs4944885 | EBV EBNA MFI (log) | -0.070 | 0.002 | <5e-08 | -0.013 | 0.017 | 0.456 |
| rs543746207 | EBV EBNA MFI (log) | 0.312 | 0.009 | <5e-08 | -0.057 | 0.068 | 0.403 |
| rs6123359 | EBV EBNA MFI (log) | -0.034 | 0.003 | <5e-08 | 0.003 | 0.027 | 0.907 |
| rs71467497 | EBV EBNA MFI (log) | 0.035 | 0.006 | <5e-08 | -0.022 | 0.050 | 0.662 |
| rs71601787 | EBV EBNA MFI (log) | -0.042 | 0.002 | <5e-08 | 0.017 | 0.017 | 0.316 |
| rs7657132 | EBV EBNA MFI (log) | 0.014 | 0.002 | <5e-08 | 0.019 | 0.017 | 0.282 |
| rs7675923 | EBV EBNA MFI (log) | -0.026 | 0.003 | <5e-08 | -0.015 | 0.022 | 0.514 |
| rs11023212 | EBV IgG Seropositivity | 0.072 | 0.002 | <5e-08 | 0.074 | 0.071 | 0.300 |
| rs117592720 | EBV IgG Seropositivity | -0.070 | 0.006 | <5e-08 | 0.016 | 0.208 | 0.938 |
| rs146128209 | EBV IgG Seropositivity | 0.058 | 0.004 | <5e-08 | -0.004 | 0.132 | 0.978 |
| rs1792287 | EBV IgG Seropositivity | -0.022 | 0.002 | <5e-08 | 0.000 | 0.076 | 0.999 |
| rs2585442 | EBV IgG Seropositivity | -0.036 | 0.002 | <5e-08 | 0.069 | 0.080 | 0.388 |
| rs2616279 | EBV IgG Seropositivity | 0.023 | 0.003 | <5e-08 | -0.003 | 0.093 | 0.977 |
| rs4694423 | EBV IgG Seropositivity | 0.101 | 0.002 | <5e-08 | -0.007 | 0.068 | 0.922 |
| rs4944885 | EBV IgG Seropositivity | -0.070 | 0.002 | <5e-08 | -0.136 | 0.071 | 0.055 |
| rs543746207 | EBV IgG Seropositivity | 0.312 | 0.009 | <5e-08 | -0.402 | 0.293 | 0.171 |
| rs6123359 | EBV IgG Seropositivity | -0.034 | 0.003 | <5e-08 | 0.073 | 0.113 | 0.518 |
| rs71467497 | EBV IgG Seropositivity | 0.035 | 0.006 | <5e-08 | 0.030 | 0.203 | 0.883 |
| rs71601787 | EBV IgG Seropositivity | -0.042 | 0.002 | <5e-08 | -0.155 | 0.072 | 0.031 |
| rs7657132 | EBV IgG Seropositivity | 0.014 | 0.002 | <5e-08 | 0.150 | 0.072 | 0.038 |
| rs7675923 | EBV IgG Seropositivity | -0.026 | 0.003 | <5e-08 | 0.009 | 0.093 | 0.922 |
| rs11023212 | EBV VCA MFI (log) | 0.072 | 0.002 | <5e-08 | -0.005 | 0.017 | 0.748 |
| rs117592720 | EBV VCA MFI (log) | -0.070 | 0.006 | <5e-08 | -0.003 | 0.050 | 0.947 |
| rs146128209 | EBV VCA MFI (log) | 0.058 | 0.004 | <5e-08 | 0.021 | 0.031 | 0.495 |
| rs1792287 | EBV VCA MFI (log) | -0.022 | 0.002 | <5e-08 | -0.031 | 0.018 | 0.079 |
| rs2585442 | EBV VCA MFI (log) | -0.036 | 0.002 | <5e-08 | -0.022 | 0.019 | 0.247 |
| rs2616279 | EBV VCA MFI (log) | 0.023 | 0.003 | <5e-08 | -0.007 | 0.022 | 0.756 |
| rs4694423 | EBV VCA MFI (log) | 0.101 | 0.002 | <5e-08 | 0.014 | 0.016 | 0.392 |
| rs4944885 | EBV VCA MFI (log) | -0.070 | 0.002 | <5e-08 | -0.029 | 0.016 | 0.079 |
| rs543746207 | EBV VCA MFI (log) | 0.312 | 0.009 | <5e-08 | -0.060 | 0.067 | 0.369 |
| rs6123359 | EBV VCA MFI (log) | -0.034 | 0.003 | <5e-08 | 0.002 | 0.026 | 0.937 |
| rs71467497 | EBV VCA MFI (log) | 0.035 | 0.006 | <5e-08 | 0.003 | 0.048 | 0.943 |
| rs71601787 | EBV VCA MFI (log) | -0.042 | 0.002 | <5e-08 | 0.008 | 0.017 | 0.611 |
| rs7657132 | EBV VCA MFI (log) | 0.014 | 0.002 | <5e-08 | -0.004 | 0.017 | 0.809 |
| rs7675923 | EBV VCA MFI (log) | -0.026 | 0.003 | <5e-08 | -0.010 | 0.022 | 0.646 |
| rs11023212 | EBV Zebra MFI (log) | 0.072 | 0.002 | <5e-08 | 0.003 | 0.017 | 0.841 |
| rs117592720 | EBV Zebra MFI (log) | -0.070 | 0.006 | <5e-08 | -0.020 | 0.050 | 0.697 |
| rs146128209 | EBV Zebra MFI (log) | 0.058 | 0.004 | <5e-08 | -0.047 | 0.031 | 0.125 |
| rs1792287 | EBV Zebra MFI (log) | -0.022 | 0.002 | <5e-08 | -0.039 | 0.018 | 0.027 |
| rs2585442 | EBV Zebra MFI (log) | -0.036 | 0.002 | <5e-08 | -0.011 | 0.019 | 0.543 |
| rs2616279 | EBV Zebra MFI (log) | 0.023 | 0.003 | <5e-08 | -0.014 | 0.022 | 0.522 |
| rs4694423 | EBV Zebra MFI (log) | 0.101 | 0.002 | <5e-08 | -0.012 | 0.016 | 0.466 |
| rs4944885 | EBV Zebra MFI (log) | -0.070 | 0.002 | <5e-08 | 0.007 | 0.017 | 0.676 |
| rs543746207 | EBV Zebra MFI (log) | 0.312 | 0.009 | <5e-08 | 0.015 | 0.067 | 0.817 |
| rs6123359 | EBV Zebra MFI (log) | -0.034 | 0.003 | <5e-08 | -0.001 | 0.026 | 0.962 |
| rs71467497 | EBV Zebra MFI (log) | 0.035 | 0.006 | <5e-08 | -0.121 | 0.048 | 0.012 |
| rs71601787 | EBV Zebra MFI (log) | -0.042 | 0.002 | <5e-08 | 0.014 | 0.017 | 0.402 |
| rs7657132 | EBV Zebra MFI (log) | 0.014 | 0.002 | <5e-08 | 0.008 | 0.017 | 0.648 |
| rs7675923 | EBV Zebra MFI (log) | -0.026 | 0.003 | <5e-08 | -0.005 | 0.022 | 0.802 |
| rs11023212 | Multiple Sclerosis | 0.072 | 0.002 | <5e-08 | -0.033 | 0.017 | 0.057 |
| rs1792287 | Multiple Sclerosis | -0.022 | 0.002 | <5e-08 | 0.010 | 0.018 | 0.594 |
| rs2616279 | Multiple Sclerosis | 0.023 | 0.003 | <5e-08 | -0.069 | 0.023 | 0.002 |
| rs4694423 | Multiple Sclerosis | 0.101 | 0.002 | <5e-08 | -0.046 | 0.019 | 0.014 |
| rs4944885 | Multiple Sclerosis | -0.070 | 0.002 | <5e-08 | 0.019 | 0.018 | 0.276 |
| rs6123359 | Multiple Sclerosis | -0.034 | 0.003 | <5e-08 | 0.044 | 0.028 | 0.115 |
| rs71467497 | Multiple Sclerosis | 0.035 | 0.006 | <5e-08 | -0.063 | 0.047 | 0.181 |
| rs71601787 | Multiple Sclerosis | -0.042 | 0.002 | <5e-08 | -0.012 | 0.019 | 0.525 |
| rs7657132 | Multiple Sclerosis | 0.014 | 0.002 | <5e-08 | 0.004 | 0.018 | 0.820 |
| rs7675923 | Multiple Sclerosis | -0.026 | 0.003 | <5e-08 | -0.010 | 0.022 | 0.663 |

| **Supplementary Table 3 – SNP level estimates for MR of vitamin D on EBV DNA load** | | | | | | | |
| --- | --- | --- | --- | --- | --- | --- | --- |
| SNP | outcome | beta.exposure | se.exposure | pval.exposure | beta.outcome | se.outcome | pval.outcome |
| rs117592720 | EBV DNA load in blood - AoU EA cohort | -0.070 | 0.006 | <5e-08 | 0.001 | 0.006 | 0.960 |
| rs146128209 | EBV DNA load in blood - AoU EA cohort | 0.058 | 0.004 | <5e-08 | -0.007 | 0.004 | 0.069 |
| rs2585442 | EBV DNA load in blood - AoU EA cohort | -0.036 | 0.002 | <5e-08 | -0.005 | 0.002 | 0.028 |
| rs2616279 | EBV DNA load in blood - AoU EA cohort | 0.023 | 0.003 | <5e-08 | 0.000 | 0.003 | 0.930 |
| rs543746207 | EBV DNA load in blood - AoU EA cohort | 0.312 | 0.009 | <5e-08 | 0.008 | 0.009 | 0.510 |
| rs6123359 | EBV DNA load in blood - AoU EA cohort | -0.034 | 0.003 | <5e-08 | 0.001 | 0.003 | 0.730 |
| rs71467497 | EBV DNA load in blood - AoU EA cohort | 0.035 | 0.006 | <5e-08 | 0.008 | 0.006 | 0.190 |
| rs117592720 | EBV DNA load in blood - UKB and AoU EA cohorts combined | -0.070 | 0.006 | <5e-08 | 0.002 | 0.003 | 0.479 |
| rs146128209 | EBV DNA load in blood - UKB and AoU EA cohorts combined | 0.058 | 0.004 | <5e-08 | 0.000 | 0.002 | 0.919 |
| rs2585442 | EBV DNA load in blood - UKB and AoU EA cohorts combined | -0.036 | 0.002 | <5e-08 | -0.002 | 0.001 | 0.155 |
| rs2616279 | EBV DNA load in blood - UKB and AoU EA cohorts combined | 0.023 | 0.003 | <5e-08 | 0.000 | 0.002 | 0.884 |
| rs543746207 | EBV DNA load in blood - UKB and AoU EA cohorts combined | 0.312 | 0.009 | <5e-08 | 0.006 | 0.005 | 0.172 |
| rs6123359 | EBV DNA load in blood - UKB and AoU EA cohorts combined | -0.034 | 0.003 | <5e-08 | -0.002 | 0.002 | 0.359 |
| rs71467497 | EBV DNA load in blood - UKB and AoU EA cohorts combined | 0.035 | 0.006 | <5e-08 | 0.000 | 0.003 | 0.871 |
| rs117592720 | EBV DNA load in blood - UKB cohort | -0.070 | 0.006 | <5e-08 | 0.002 | 0.004 | 0.450 |
| rs146128209 | EBV DNA load in blood - UKB cohort | 0.058 | 0.004 | <5e-08 | 0.003 | 0.003 | 0.270 |
| rs2585442 | EBV DNA load in blood - UKB cohort | -0.036 | 0.002 | <5e-08 | 0.000 | 0.002 | 0.740 |
| rs2616279 | EBV DNA load in blood - UKB cohort | 0.023 | 0.003 | <5e-08 | 0.000 | 0.002 | 0.850 |
| rs543746207 | EBV DNA load in blood - UKB cohort | 0.312 | 0.009 | <5e-08 | 0.006 | 0.006 | 0.200 |
| rs6123359 | EBV DNA load in blood - UKB cohort | -0.034 | 0.003 | <5e-08 | -0.003 | 0.002 | 0.170 |
| rs71467497 | EBV DNA load in blood - UKB cohort | 0.035 | 0.006 | <5e-08 | -0.002 | 0.004 | 0.550 |
| rs117592720 | EBV DNA load in saliva - AoU EA cohort | -0.070 | 0.006 | <5e-08 | 0.026 | 0.035 | 0.450 |
| rs146128209 | EBV DNA load in saliva - AoU EA cohort | 0.058 | 0.004 | <5e-08 | -0.021 | 0.021 | 0.320 |
| rs2585442 | EBV DNA load in saliva - AoU EA cohort | -0.036 | 0.002 | <5e-08 | 0.003 | 0.013 | 0.810 |
| rs2616279 | EBV DNA load in saliva - AoU EA cohort | 0.023 | 0.003 | <5e-08 | 0.003 | 0.015 | 0.840 |
| rs543746207 | EBV DNA load in saliva - AoU EA cohort | 0.312 | 0.009 | <5e-08 | 0.049 | 0.053 | 0.350 |
| rs6123359 | EBV DNA load in saliva - AoU EA cohort | -0.034 | 0.003 | <5e-08 | 0.007 | 0.018 | 0.690 |
| rs71467497 | EBV DNA load in saliva - AoU EA cohort | 0.035 | 0.006 | <5e-08 | 0.013 | 0.033 | 0.700 |

| **Supplementary Table 4 – Test of Heterogeneity** | | | | |
| --- | --- | --- | --- | --- |
| **Outcome** | **Exposure** | **Q** | **Qdf (degrees of freedom)** | **Q p-value** |
| EBV EA-D MFI (log) | Vitamin D Concentration | 27.09 | 16 | 0.04 |
| EBV IgG Seropositivity |  | 18.09 | 16 | 0.32 |
| EBV Zebra MFI (log) |  | 27.24 | 16 | 0.04 |
| EBV EBNA MFI (log) |  | 10.41 | 16 | 0.84 |
| Multiple Sclerosis |  | 14.49 | 12 | 0.27 |
| EBV VCA MFI (log) |  | 13.20 | 16 | 0.66 |
| EBV DNA load in saliva - AoU EA cohort | Vitamin D Concentration | 2.98 | 7 | 0.89 |
| EBV DNA load in blood - UKB cohort |  | 3.42 | 7 | 0.84 |
| EBV DNA load in blood - AoU EA cohort |  | 12.00 | 7 | 0.10 |
| EBV DNA load in blood - UKB and AoU EA cohorts combined |  | 2.73 | 7 | 0.91 |

| **Supplementary Table 5 – Test of Heterogeneity** | | | | |
| --- | --- | --- | --- | --- |
| **Outcome** | **Exposure** | **Egger intercept** | **Standard error** | **P-value** |
| EBV EA-D MFI (log) | Vitamin D Concentration | 0 | 0.012 | 0.98 |
| EBV IgG Seropositivity |  | 0.07 | 0.039 | 0.11 |
| EBV Zebra MFI (log) |  | 0.01 | 0.012 | 0.43 |
| EBV EBNA MFI (log) |  | 0.01 | 0.009 | 0.19 |
| Multiple Sclerosis |  | -0.005 | 0.013 | 0.71 |
| EBV VCA MFI (log) |  | 0.015 | 0.009 | 0.12 |
| EBV DNA load in saliva - AoU EA cohort | Vitamin D Concentration | -0.008 | 0.011 | 0.50 |
| EBV DNA load in blood - UKB cohort |  | 0 | 0.001 | 0.83 |
| EBV DNA load in blood - AoU EA cohort |  | 0.001 | 0.003 | 0.69 |
